# Antibody responses to SARS-CoV2 are distinct in children with MIS-C compared to adults with COVID-19

**DOI:** 10.1101/2020.07.12.20151068

**Authors:** Stuart P. Weisberg, Thomas Connors, Yun Zhu, Matthew Baldwin, Wen-hsuan Lin, Sandeep Wontakal, Peter A. Szabo, Steven B. Wells, Pranay Dogra, Joshua Gray, Emma Idzikowski, Francesca T. Bovier, Julia Davis-Porada, Rei Matsumoto, Maya Meimei Li Poon, Michael Chait, Cyrille Mathieu, Branka Horvat, Didier Decimo, Zachary C. Bitan, Francesca La Carpia, Stephen A. Ferrara, Emily Mace, Joshua Milner, Anne Moscona, Eldad Hod, Matteo Porotto, Donna L. Farber

## Abstract

Clinical manifestations of COVID-19 caused by the novel coronavirus SARS-CoV-2 are associated with age. While children are largely spared from severe respiratory disease, they can present with a SARS-CoV-2-associated multisystem inflammatory syndrome (MIS-C) similar to Kawasaki’s disease. Here, we show distinct antibody (Ab) responses in children with MIS-C compared to adults with severe COVID-19 causing acute respiratory distress syndrome (ARDS), and those who recovered from mild disease. There was a reduced breadth and specificity of anti-SARS-CoV-2-specific antibodies in MIS-C patients compared to the COVID patient groups; MIS-C predominantly generated IgG Abs specific for the Spike (S) protein but not for the nucleocapsid (N) protein, while the COVID-19 cohorts had anti-S IgG, IgM and IgA Abs, as well as anti-N IgG Abs. Moreover, MIS-C patients had reduced neutralizing activity compared to both COVID-19 cohorts, indicating a reduced protective serological response. These results suggest a distinct infection course and immune response in children and adults who develop severe disease, with implications for optimizing treatments based on symptom and age.

## Main text

A new clinical manifestation of SARS-CoV-2 infection has been identified in children, designated Multisystem Inflammatory Syndrome in Children (MIS-C), characterized by a systemic inflammatory response with similarities to Kawasaki disease and toxic shock syndrome but lacking respiratory illness that is a hallmark of COVID-19 in adults^1,2^. The nature of the immune response in MIS-C relative to the more common forms of COVID-19 is not well understood. The generation of virus-specific antibodies which neutralize or block infectivity is the most consistent correlate of protective immunity for multiple infections and vaccines^3,4^. Antibodies specific for the Spike (S) protein of SARS-Cov-2 which binds the cellular receptor for viral entry, have been detected in actively infected patients and in patients with mild disease who recovered^5-8^. Anti-S antibodies can exhibit neutralizing activity and are currently being pursued as a therapeutic option for infusion into patients during severe disease and for targeted generation in vaccines^9-11 12^.

In this study, we investigated the specificity and functionality of the antibody response and its protective capacity in three patient cohorts seen at New York Presbyterian hospital during the height of the pandemic from March-June, 2020 ^1,10,13,14^, including patients with mild COVID-19 respiratory disease who recovered and were recruited as convalescent plasma donors (CPD), hospitalized patients with the severest form of respiratory disease (COVID-ARDS), and a cohort of children hospitalized with MIS-C (See Table 1 for clinical characteristics). The COVID-19 patient cohorts represented a broad age range (14-84 years) while MIS-C ages were younger (4-17yrs) (Table 1). Co-morbidities were relatively common in adult subjects with COVID-ARDS, but not in MIS-C. Samples were obtained from hospitalized patients within 24-36hrs of being admitted or intubated, which represented a shorter time post-symptom onset in MIS-C compared to COVID ARDS patients, respectively. Both MIS-C and COVID-ARDS subjects exhibited markers of systemic inflammation including highly elevated levels of IL-6 and C-reactive protein (CRP), while ferritin and lactate dehydrogenase (LDH), were significantly increased in COVID-ARDS compared to MIS-C subjects (Table 1). None of the MIS-C subjects developed respiratory failure or ARDS, and had reduced overall organ damage when compared to the COVID ARDS group, indicating distinct inflammatory and systemic responses in these two disease manifestations.

**Table 1.** Demographic and Clinical Data.

|  | CPD (n=19) | COVID-ARDS (n=14) | MIS-C (n=15) | P value |
| --- | --- | --- | --- | --- |
| **Clinical Characteristics** |  |  |  |  |
| Age, years median (range) | 45 (28-69) | 56 (14-84) | 11 (4-17) |  |
| Sex, male (%) | 10 (53%) | 11 (79%) | 7 (47%) |  |
| Body Mass Index, kg/m<sup>2</sup> median (IQR) | na | 33.9 (28.4-36.6) | 18.9 (17.3-24.7) |  |
| SOFA Score<sup>a</sup>, median (IQR)<sup>b</sup> | na | 11 (10-14) | 4 (3-7) |  |
| ARDS (%) | na | 14 (100%) | 0 (0%) |  |
| In-hospital mortality (%) | na | 6 (43)%<sup>c</sup> | 0 (0%) |  |
| SARS-CoV-2 PCR Positive, n (%) | na | 14 (100%) | 7 (47%)<sup>d</sup> |  |
| Symptom Duration, days median (IQR)<sup>e</sup> | 24 (19-37) | 17 (14-20) | 6 (4-7) |  |
| **Race or ethnic group (%)<sup>f</sup>** |  |  |  |  |
| Hispanic or Latino | 1 (5%) | 5 (36%) | 3 (20%) |  |
| Black or African American | 0 | 3 (21%) | 7 (47%) |  |
| White | 10 (53%) | 3 (21%) | 6 (40%) |  |
| Asian | 6 (32%) | 0 | 0 |  |
| Pacific Islander | 2 (11%) | 0 | 0 |  |
| Other or Unknown | 1 (5%) | 5 (36%) | 1 (7%) |  |
| **Co-Morbidities (%)<sup>f</sup>** |  |  |  |  |
| Hypertension | na | 4 (29%) | 0 |  |
| Diabetes | na | 4 (29%) | 0 |  |
| Current or Former Smoker | na | 3 (21%) | 0 |  |
| COPD or ILD | na | 2 (14%) | 0 |  |
| Chronic Neurologic Diseases or Dementia | na | 1 (7%) | 0 |  |
| Asthma | na | 1 (7%) | 3 (23%) |  |
| **Treatment** |  |  |  |  |
| Convalescent Plasma | na | 4(29%) | 0 |  |
| Hydrocortisone | na | 6 (40%) | 3 (20%) |  |
| Intravenous Immunoglobulin | na | 1 (7%) | 13 (87%) |  |
| Methylprednisone | na | 11 (73%) | 14 (93%) |  |
| Monoclonal Antibody<sup>g</sup> | na | 7 (47%) | 2 (13%) |  |
| Remdesivir | na | 6 (40%) | 1 (7%) |  |
| **Laboratory Results (IQR)<sup>b</sup>** |  |  |  |  |
| Absolute Neutrophil Count, x10(3)/μL | na | 17.0 (9.2-29.0) | 8.8 (7.0-12.5) | 0.046 |
| Absolute Lymphocyte Count x10(3)/μL | na | 0.8 (0.6-1.4) | 0.8 (0.4-1.6) | 0.95 |
| D-dimer, μg/mL<sup>h</sup> | na | 7.3 (2.0-14.5) | 2.9 (1.5-4.5) | 0.09 |
| Ferritin, ng/mL<sup>h</sup> | na | 1947 (1010-2880) | 496 (278-694) | 0.0002 |
| High Sensitivity CRP, mg/L<sup>h</sup> | na | 132 (80-241) | 204 (49-300) | 0.78 |
| Interleukin-6, pg/mL<sup>h</sup> | na | 89 (61-315) | 307 (132-315) | 0.63 |
| Lactate Dehydrogenase, U/L<sup>h</sup> | na | 776 (638-1379) | 268 (229-373) | 0.0003 |
| Procalcitonin, ng/mL | na | 0.38 (0.28-3.3) | 5.8 (1.92-33.3) | 0.013 |
| Troponin T, high sensitivity, ng/L | na | 24 (17-67) | 19 (6-88) | 0.62 |
| Abbreviations: CPD, convalescent plasma donor; ARDS, Acute Respiratory Distress Syndrome; MIS-C Multisystem Inflammatory Syndrome in Children; SOFA, Sequential Organ Failure Assessment; IQR, Interquartile Range; PCR, Polymerase chain reaction; CRP, C-Reactive Protein; COPD, Chronic Obstructive Pulmonary Disease; ILD, Interstitial Lung Disease |  |  |  |  |
| a Pediatric and Adult specific scoring applied to groups; not meant for direct comparison |  |  |  |  |
| b Day of admission for MIS-C and day of intubation for COVID-ARDS |  |  |  |  |
| c 30 Day In-hospital mortality, 4 patients remain hospitalized |  |  |  |  |
| d Indeterminate tests were treated as positive |  |  |  |  |
| e Pre-study symptom duration; respiratory symptoms for CPD/ARDS groups and symptoms of MIS-C for MIS-C group |  |  |  |  |
| f Individuals included in all groups for which they identified/met criteria |  |  |  |  |
| g Received or enrolled in blinded trial; Anikinra (n=2 MIS-C), Tocilizumab (n=5 COVID-ARDS), Sarilumab (n=2 COVID ARDS) |  |  |  |  |
| h Values above upper limit entered as; D-Dimer (20 mg/mL), Ferritin (100,000 ng/mL), CRP (200 mg/L), Interleukin-6 (315 pg/mL), Lactate Dehydrogenase (5000 U/L) |  |  |  |  |

We initially established that MIS-C and COVID-19 in our patients are associated with a specific immune response to SARS-CoV-2, by testing binding of plasma IgG to recombinant S proteins expressed on the cell surface. Plasma from all patient samples but not pre-pandemic control samples bound SARS-CoV-2 S protein and the common circulating D614G S protein variant^15^ but not to S protein from SARS-CoV-1 or MERS coronaviruses (Extended Data, Fig. 1). We then quantitated the Ab response in each cohort in terms of specificity and Ab class, including IgM generated initially in a primary response and IgG and IgA classes prominent in serum and secretions, respectively. Anti-S antibodies of the IgM, IgG and IgA classes were significantly elevated in COVID and CPD donors relative to negative control plasma obtained from pre-pandemic samples; COVID-ARDS patients had the highest levels of anti-S Abs for all classes (Fig. 1a-c). By contrast, anti-S antibodies in patients with MIS-C were predominantly IgG and to a lesser extent IgA—both at similar levels as the CPD donors while anti-S IgM levels in MIS-C were at the same low level as negative control plasma (Fig. 1a-c, left). Moreover, the ratio of anti-S IgG to IgM was significantly higher in MIS-C patients compared to the COVID-ARDS and CPD subjects (MIS-C anti-S IgG/IgM ratio 3.35 vs COVID ARDS 1.48, p<0.0001 and vs CPD 1.95, p=0.001) indicating skewed production of anti-S IgG in MIS-C. No significant correlation was found between any of the anti-S Ig isotypes and patient age (Fig. 1a-c, right).

**Figure 1.**
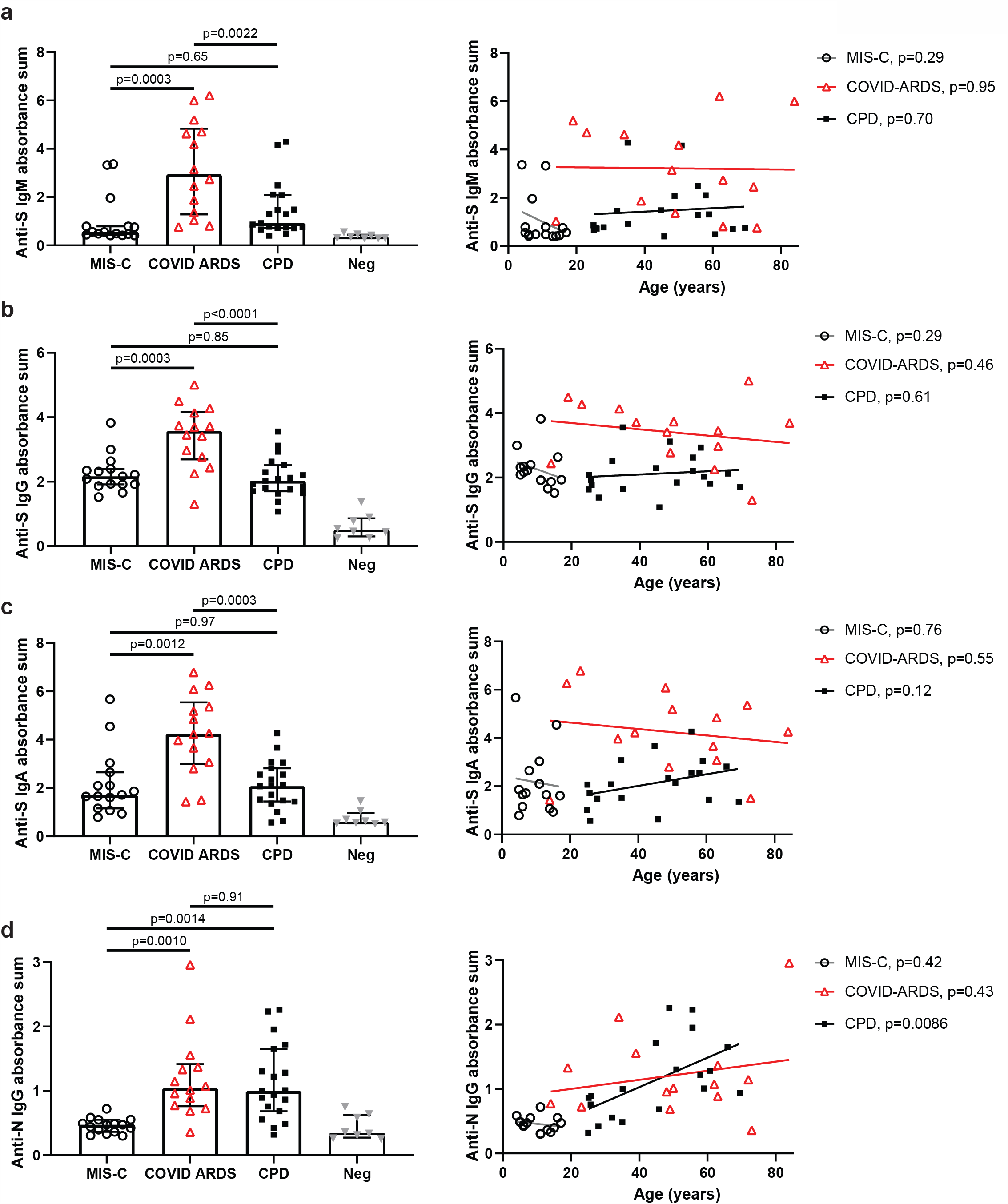
Distinct SARS-CoV-2 antibody responses in MIS-C compared to severe and mild COVID-19 disease. Levels of antibodies to SARS-CoV-2 spike (S) and nucleocapsid (N) proteins were assessed using serial dilutions of patient plasma in an indirect ELISA assay to detect anti-S IgM (a), anti-S IgG (b). anti-S IgA (c) and anti-N IgG (d). Shown (left) is the absorbance sum across 6 serial 1:4 plasma dilutions from patients with COVID-19-related Multisystem Inflammatory Syndrome in Children (MIS-C, white circles, n=15); patients with COVID-19 induced acute respiratory distress syndrome (COVID ARDS, red triangles, n=14) convalescent plasma donors (CPD, black squares, n=19), and control plasma from pre-pandemic donors (grey triangles; Neg, n=8). ±IQR. P value as calculated by one-way ANOVA with Tukey’s multiple comparisons test. Shown (right) are subject antibody levels plotted against patient age with the best fit line and p value as calculated using simple linear regression.

We also assessed the presence of antibodies specific for the SARS-CoV-2 nucleocapsid (N) protein, which complexes with viral RNA, and is involved in viral replication during active infection^16^. The MIS-C group showed significantly lower anti-N IgG levels compared to both the COVID ARDS and CPD groups (Fig. 1d, left). In addition, a significant correlation of anti-N IgG levels with subject age was observed within the CPD group but not the COVID ARDS and MIS-C groups (Fig. 1d, right). These results show that MIS-C patients generate mostly anti-S IgG antibodies, while COVID-19 adult patient groups had a broader antibody response in terms of isotype and specificities with COVID ARDS patients exhibiting the highest titers and antibody levels of all patient groups.

The ability of antibodies to provide protection is correlated with their neutralizing activity in blocking virus infection. We developed a cell-based pseudovirus assay based on a system previously reported^17,18^ in which multi-cycle infection of RFP-expressing Vesicular stomatitis virus (VSV) pseudotyped with SARS-CoV-2 S protein is measured in the presence of serially diluted plasma samples (see methods). We validated this assay in two ways: First, we compared neutralizing activity of plasma samples tested in the pseudovirus assay to activity measured in live virus microneutralization assay based on inhibition of cytopathic effect^19^, and found a direct correlation in neutralizing activity calculated from the pseudovirus and live virus assay over a wide range of neutralizing activity (r^2^=0.54, slope=0.95, p=0.0044) (Extended Data Fig. 2a). Second, we compared neutralizing activity of samples with known anti-S IgG positivity by ELISA (n=18) to samples that tested negative (n=30), and found significantly increased neutralizing activity in the ELISA-positive compared to ELISA-negative samples (Fig. S1), demonstrating both specificity and sensitivity of the SARS-CoV-2 pseudovirus assay.

We measured SARS-CoV-2 neutralizing activity in the different patient groups using the pseudovirus assay. Overall, the CPD and COVID ARDS plasma samples exhibited greater neutralizing activity compared to plasma from MIS-C patients (Fig. 2a); plasma from COVID ARDS patients show the highest neutralizing potency of the three groups across the dilution series (Extended Data Fig. 2b). Within the MIS-C group, 4/15 (26.7%) patients had neutralizing activity > 2 SD above the mean of negative controls compared to 11/19 (57.9%) in the CPD group and 13/14 (92.9%) in the COVID ARDS group. These variations in neutralizing activity between groups did not correlate with patient age (Fig. 2b). MIS-C patients also maintained the same levels of anti-S IgG and neutralizing activity 2-4 weeks after hospital discharge based on paired analysis of the follow-up compared to the retested primary sample in 10/15 (66.7%) of the patients (Fig. 2c). These results suggest that the distinct SARS-CoV-2 antibody profile in MIS-C remains stable during recovery.

**Figure 2.**
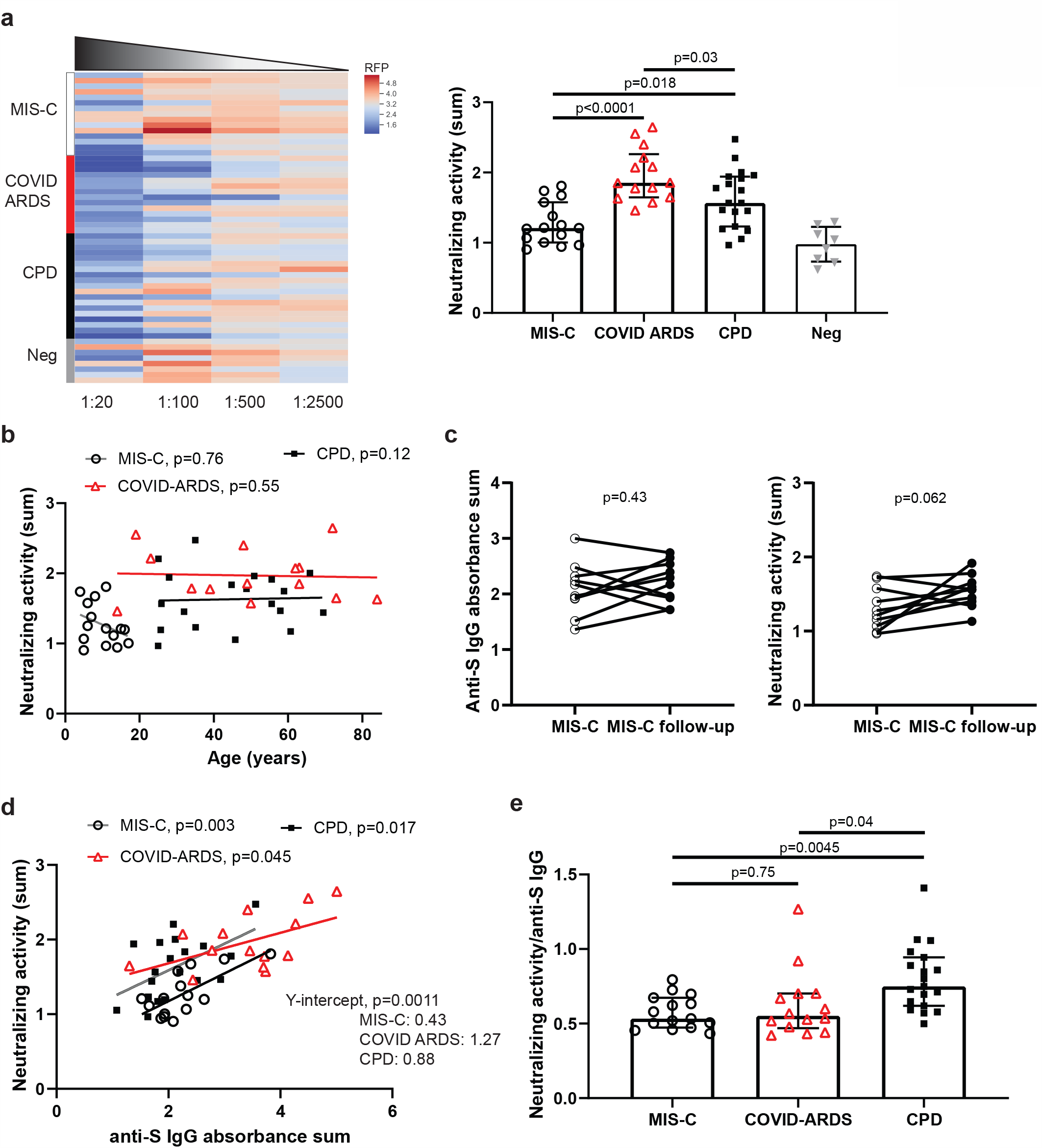
Reduced SARS-CoV-2 neutralizing activity associated with MIS-C compared to severe and mild disease independent of age. a, b Neutralizing activity for SARS-CoV-2-specific antibodies was determined using a pseudovirus assay in which multi-cycle replication of SARS-CoV-2 S protein and red fluorescent protein (RFP)-expressing viruses was tested in the presence of 4 serial 1:5 dilutions of patient plasma. (see methods). For each sample, neutralizing activity is calculated based on the inhibition of RFP signal across dilutions. Shown is a heat map (a, left) of the normalized RFP signal across plasma titrations from patients with MIS-C (white bar, n=15); COVID ARDS (red bar, n=13), convalescent plasma donors (CPD, black bar, n=19), and control plasma from pre-pandemic donors (grey bar; Neg, n=8). The sum of the reciprocal values at the dilutions are shown (a, right) compiled from each of the patient and control groups. ±IQR. P value as calculated by one-way ANOVA with Tukey’s multiple comparisons test. These values are also plotted against patient age (b) with the best fit line and P values calculated using simple linear regression. c, The neutralizing activity and anti-S IgG levels of MIS-C patients (n=10) during the acute phase of illness and at a follow-up visit 2-4 weeks after hospital discharge. P value as calculated by paired t-test. d, The anti-S IgG absorbance sums were plotted against SARS-CoV-2 neutralizing activity from patients with MIS-C (white circles, n=15, r^2^=0.5); COVID ARDS (red triangles, n=13,, r^2^=0.29) and CPD (black squares, n=19, r^2^=0.29). Best fit lines, r^2^values, P values and Y-intercepts as calculated by simple linear regression. e, The ratio of the neutralizing activity to anti-S IgG level for patients in each group is compiled. ±IQR. P value as calculated by one-way ANOVA with Tukey’s multiple comparisons test.

Only a small fraction of antibodies raised against viral antigens will have neutralizing activity against the virus, which correlates with protective capacity^20^. By linear regression, there was significant correlation between the level of anti-S IgG and neutralizing activity within each group, albeit with a lower elevation and y intercept for MIS-C group relative to the COVID-ARDS and CPD groups (Fig. 2d). Comparing the ratio of plasma neutralizing activity to anti-S IgG level (Neu:IgG) for each sample reveals a significantly higher Neu:IgG for the CPD group compared to the MIS-C and COVID ARDS groups Fig. 2e). This analysis suggests that the anti-SARS-CoV-2 immune response in the most critically ill patients was less targeted to protection. It remains to be determined whether the distinct serological responses observed in the MIS-C patients is a characteristic of the pediatric immune response to SARS-CoV-2 or specific for this syndrome. Plasma samples from a large cohort of otherwise healthy children who recovered uneventfully from SARS-CoV-2 infection could not be obtained due to restrictions on clinical research needed to slow the spread of the pandemic. Comparison of samples from 3 pediatric COVID, non-MIS-C patients - two of which had pre-existing hematologic or oncologic disease - showed similar anti-S IgG and neutralizing activity to the follow-up samples from recovered MIS-C patients (Table S1).

Our results show clear quantitative and qualitative differences in the functional SARS-CoV-2-specific antibody response in children with MIS-C compared to adults with mild and severe COVID-19. Children with MIS-C showed the least extensive antibody response that was largely limited to IgG anti-S antibodies with the lowest overall level of neutralizing activity, while patients with the most severe form of COVID ARDS had the highest overall levels and breadth of anti-SARS-CoV2 antibodies and the highest overall neutralizing activity. Despite MIS-C patients having the shortest interval among the groups between symptom onset and sample collection, their SARS-CoV-2-specific antibody response is strongly biased towards IgG vs IgM production, suggesting that MIS-C may arise during a late stage after initial infection.

SARS-CoV-2 is a novel virus and as such, both adults and children lack adaptive immunologic memory and are experiencing primary exposures. The less robust antibody response in children with MIS-C could result from a less productive infection in children, also supported by the lack of anti-N-specific antibodies in MIS-C that requires lysis of virally infected cells. Interestingly, we found that among the CPD group, there was an age-dependence in production of anti-N antibodies, indicating that reduced anti-N antibodies may be a feature of infection in younger individuals. While a small cohort of non-MIS-C patients showed a similar Ab response as MIS-C, studies in larger cohorts of healthy children who were exposed to or developed COVID-19 are needed to assess the pediatric immune response to SARS-CoV-2. The presence of SARS-CoV-2-specific T cells in the peripheral blood of recovered and COVID-ARDS adult patients has been recently reported^21,22^, though the protective capacity of these T cells is unclear. For respiratory viruses, T cells which localize and establish residency in the lung mediate protective immunity, as shown in mouse models^23-25^. Because younger individuals have increased number of naïve T cells in different sites to respond to new pathogens^26^, it is possible that a robust T cell response efficaciously clears infection in the lung preventing severe respiratory disease in children, and a low level, persistent infection in other sites may build up over time in some children, resulting in MIS-C. Other direct effects of SARS-CoV-2 on immune activation has also been proposed ^27^.

Current therapeutic options for severe COVID-19 disease have included convalescent plasma, anti-inflammatory immune modulators, and antivirals. The mainstay of treatment for MIS-C subjects has been steroids and intravenous immunoglobulin (IVIG) infusions, largely deriving from experience in children with Kawasaki syndrome^1,2^. Our results showing inefficient neutralizing activity of the MIS-C antibody response suggest that targeted treatment of these children with functionally neutralizing anti-SARS-CoV-2 neutralizing products should be considered as a therapeutic option in MIS-C^28,29^. The continued pursuit of additional therapeutic modalities remains crucial while awaiting an effective vaccine.

## METHODS

### Subjects

We recruited a total of 48 subjects from the Morgan Stanley Children’s Hospital of New York and Columbia University Irving Medical Center/New York Presbyterian Hospital (CUIMC/NYP) who represented 3 distinct clinical manifestations of SARS-CoV-2 infection. Individuals (n=19) donating blood as part of our institution’s convalescent plasma trial (convalescent plasma donors, CPD) following a history of recent illness consistent with SARS-CoV-2 infection and subsequently found to have SARS-CoV-2 antibody positive serology. Patients with severe COVID-19 and ARDS (n=14) who tested positive for SARS-CoV-2 from nasopharyngeal swabs. Pediatric patient with MIS-C (n=15) and confirmed SARS-CoV-2 antibody positive serology. ARDS was defined by clinical consensus criteria; including infiltrates on chest radiograph and a PaO2/FiO2 ratio of less than 300, or pediatric criteria equivalent ^30,31^. MIS-C was defined using the Center for Disease Control definition; <21 years of age, fever >38°C for >24 hours, laboratory evidence of inflammation, hospital admission, multisystem involvement, no alternative plausible diagnosis, and positive SARS-Cov-2 serology^32^. Subjects less than 21 years of age recruited in the inpatient setting were assigned to COVID-ARDS vs MIS-C group following review of their charts by specialists in adult and pediatric critical care medicine. Sequential Organ Failure Assessment (SOFA) scores were calculated on all subjects using previously validated adult and pediatric score tools to provide additional clinical insight into subject disease severity^33-35^. This study was approved by the Institutional Review Board at Columbia University Irving Medical Center. Written consent was obtained from CPD subjects. Due to the limitations placed on direct contact with infected subjects and a need to conserve personal protective equipment, verbal informed consent was obtained from surrogates of critically ill COVID-ARDS subjects and verbal parental consent was obtained for MIS-C subjects.

### Sample Processing

Blood samples were obtained at time of outpatient donation for CPD subjects, at time of admission for MIS-C subjects, and following diagnosis of ARDS for COVID-ARDS patients. Plasma was isolated from whole blood via centrifugation. Aliquots were frozen at −80°C prior to analysis.

### Cell-based Assay for viral Spike protein reactivity

HEK293T cells were transfected with full length, codon optimized S-protein from SARS-CoV-2, SARS-CoV-2 D614G variant, MERS, SARS-CoV-1 or empty pCAGGS control vector (Epoch Life Science, Sugar Land, TX). Transfected cells were seeded onto 96 well plates (40,000 cells/well), cultured at 37°C, 5% CO_2_ overnight, and heat-inactivated plasma samples were added for 30 minutes on ice, followed by washing with cold PBS and fixation in 4% paraformaldehyde. The fixed cells were then stained with protein G Alexa Fluor 488 (ThermoFisher, Waltham, MA) and 4′,6-diamidino-2-phenylindole (DAPI) followed by fluorescence image acquisition using the IN Cell Analyzer 2500 HS high content analysis imaging system (Cytiva, Marlborough, MA). Image analysis was performed using ImageJ ^36^.

### Purification of SARS-CoV2 viral proteins

The ectodomain of the SARS-CoV-2 spike trimer^37^ was cloned into mammalian expression vector pCAGGS (Addgene, Watertown, MA), with a foldon tag followed by 6xHis tag and Strep tag II at the C-terminal. This expression vector was transiently transfected into HEK293F cells and the spike trimer secreted in the supernatant was purified 3-5 days post transfection by metal affinity chromatography using an Ni-NTA (Qiagen) column. SARS-CoV-2 nucleocapsid protein (N) was cloned into pET28a(+) vector (Millipore-Sigma, Burlington, MA) with an AAALE linker and 6xHis tag at the C-terminal. The NP construct was then used to transform into E. coli BL21 (DE3) pLysS cells and the target protein was produced and purified from the bacterial lysate by metal affinity chromatography using an Ni-NTA (Qiagen) column, followed by size-exclusion chromatography on a Superdex 200 10/300 GL column.

### Enzyme-linked immunosorbent assay (ELISA) for detection of virus-specific antibodies

SARS-CoV-2 spike trimer and N were coated on 96 well ELISA plates at 4°C overnight, and unbound proteins were then removed washing with PBS, following by blocking with PBS/3% non-fat dry milk. Plasma samples were serially diluted in PBST (0.1% Tween-20 in PBS) + 10% bovine calf serum starting with 1:100, and five successive four-fold dilutions into each well of the coated plate which was incubated at 37°C for 1hr, followed by washing 6 times with PBST. Peroxidase affiniPure goat anti-human IgG (H+L) antibody, anti-human IgM antibody (Jackson Immune Research, New Grove, PA), or anti-human IgA antibody (Thermofisher, Waltham, MA) was subsequently added into each well and incubated for 1hr at 37°C, washed and TMB substrate (Sigma, St. Louis) was added and the reaction was stopped using 1M sulfuric acid. Absorbance was measured at 450 nm and expressed as an optical density, or OD_450_ value. Identical serial dilutions were performed for all samples with no missing titrations.

### Pseudovirus neutralization assay

We adapted a pseudovirus-based neutralization strategy we previously developed to measure inhibition of infection by high biocontainment enveloped viruses in a large number of samples under low-level biocontainment ^17,18^. For this assay, SARS-CoV-2 S protein is pseudotyped onto recombinant vesicular stomatitis virus (VSV) that expresses red fluorescent protein (RFP) but does not express the VSV attachment protein, G (VSV-ΔG-RFP). Initially, VSV-ΔG-RFP pseudotyped with VSV G is used to infect 293T (human kidney epithelial) cells that were co-transfected with full length codon optimized SARS-CoV-2 S-protein (Epoch Life Science, Sugar Land, TX), the viral entry receptor ACE2 (Epoch Life Science, Sugar Land, TX) and green fluorescent protein (GFP). Infected HEK293T cells are then mixed at a 2 to 1 ratio with Vero (African green monkey kidney) cells which have high endogenous expression of ACE2^38^. The cells are then combined with diluted serum or plasma in 96 well plates. During the assay, infected, S protein-expressing HEK293T cells generate VSV-ΔG-RFP viruses that bear S protein which subsequently infects and drives RFP expression in Vero cells and undergo multiple cycles of entry and budding in the HEK293T cells due to the co-expression of S protein with ACE2. The GFP and RFP signals are measured 24-48hrs after plating (Infinite M1000 PRO microplate reader, Tecan, Männedorf, Switzerland), resulting in robust amplification of the S protein pseudovirus-driven RFP signal between 24-48hr. Inhibition of RFP signal amplification indicates S protein neutralizing activity in patient plasma (Figure S1). Identical, five-fold serial dilutions were performed for all samples and there were no missing titration data points for any of the samples.

### SARS-CoV-2 viral stock production

SARS-CoV-2 (2019-nCoV/USA_WA1/2020) was kindly provided to BH by World Reference Center for Emerging Viruses and Arboviruses (WRCEVA) (Galveston Texas, USA). To generate virus stocks, Vero E6 cells (kindly provided by Dr. F.-L. Cosset) were inoculated with virus at a MOI of 0.01. The virus-containing medium was harvested at 72hrs post infection, clarified by low-speed centrifugation, aliquoted, and stored at −80°C. Virus stock was quantified by limiting dilution plaque assay on Vero E6 cells as described^39,40^.

### Live virus neutralization assay

Two-fold dilutions of plasma in 50µL of Dulbecco’s Modified Eagle Media (DMEM) were incubated with 200 plaque forming unit (pfu) of SARS-CoV-2 in 50µL of DMEM for 30min at 4°C. 100µL of DMEM 4%FBS containing 4×10^4^ Vero E6 cells were added on the top of the former mix in order to have final dilution of sera from 1:50 to 1:6400 (4 wells per dilution). Cells were then incubated for 3 days at 37°C, 5% CO_2_. Cytopathic effect was revealed by crystal violet staining, and scored by an observer blinded to the study design and sample identity. Neutralization end point titers were expressed as the value of the last serum dilution that completely inhibited virus-induced cytopathic effect.

### Quantitation of antibody titrations in ELISA and neutralization assays

For quantitation of neutralization titers in the pseudovirus assay, RFP signal driven by the pseudovirus normalized to the GFP signal derived from the SARS-Cov-2 S protein and ACE2 transfected cells was measured at 24 and 48hr; the ratio of normalized RFP at 48 hours (RFP48) to normalized RFP at 24 hours (RFP24) was calculated. This ratio provides a read-out of multicycle infection of the S protein/ACE2 transfected cell monolayer by S protein-bearing pseudoviruses. Heatmaps of this ratio at all titrations for all samples were generated using the Python data visualization library, seaborn^41^. Neutralizing activity for each sample was calculated by taking the sum of the reciprocal of the RFP48/RFP24 ratio at all 6 plasma dilutions for each sample as described^42^, and also by percent inhibition of multicycle replication at each dilution calculated based on the RFP48/RFP24 ratio of the sample, control wells of maximal multicycle replication without inhibition (MAX) and control wells with 100% inhibition of multicycle replication using a lipidated SARS-CoV-2 derived peptide (MIN)^40,43,44^. The equation for % inhibition of multicycle replication: 100 X (1-(sample-MAX)/(MAX – MIN)).

### Statistical analysis

All statistical analysis was performed using Prism 8 software (GraphPad, San Diego, CA). Comparisons of clinical data between groups was performed using the Mann–Whitney U test. Comparisons of antibody levels and neutralization activity were performed using one-way Analysis of Variance (ANOVA) and Tukey’s multiple comparisons test. Correlation analysis was performed using simple linear regression.

## Data Availability

The raw data analyzed for this study are available from the authors upon request.

## ACKNOWLEDGEMENTS

We wish to express our gratitude to the Medical ICU nurse champions, Cora Garcellano, Tenzin Drukdak, Harriet Avila Raymundo, Lori Wagner, and Ricky Lee, who led the efforts to obtain patient samples for the adult ARDS patients, to Evelyn Hernandez and Lorena Gomez for their roles as clinical coordinators, and to the nurses and clinical staff in the pediatric intensive care unit of Morgan Stanley Children’s hospital. We acknowledge the dedication, commitment, and sacrifice of the other nurses, providers, and personnel who helped care for these patients during the COVID-19 crisis. We acknowledge the suffering and loss of our COVID-19 patients and of their families and our community.

## AUTHOR CONTRIBUTIONS

All authors meet authorship criteria and approve of publication. S.P.W., T.C., M.P. and D.L.F., A.M., and E.H. conceived and designed the study and assays, wrote and/or edited the paper. T.C., M.B., D.L.F., E.M., J.M., E.I., coordinated sample acquisition, recruited and consented MIS-C and COVID-ARDS patients. T.C., J.D., S.P.W., E.M., J.M. and M.B. performed compilation and analysis of clinical data from MIS-C and COVID-ARDS patients. E.H., and Z.C.B. recruited and consented convalescent plasma donors. S.P.W. and Z.C.B. performed compilation and analysis of data from the convalescent plasma study. F.L.C., S.A.F, M.P.C., and S.P.W. performed collection, isolation and storage of samples from convalescent plasma donors. P.A.S, S.B.W., P.D., J.G., E.I., R.M., and M.M.P. performed collection, isolation and storage of samples from COVID ARDS patients. E.H., S.W. and W.L. established and performed ELISA assays for quantification of anti-SARS-CoV-2 S and N protein IgG, IgM, and IgA from human serum and plasma. M.P., A.M., F.T.B and Y.Z. established and performed SARS-CoV-2 S protein pseudovirus neutralization assays. C.M., B.H., and D.D. established and performed the SARS-CoV-2 live virus neutralization assay. S.P.W., D.L.F and T.C. performed compilation and analysis of data from the ELISA, pseudovirus neutralization and live virus neutralization assays.

### Data availability statement

The raw data analyzed for this study and presented in all figures and tables are provided in Source Data. Additional supporting data are available from the authors upon request.

### Code availability statement

N/A

---

**Extended data figure 1.**
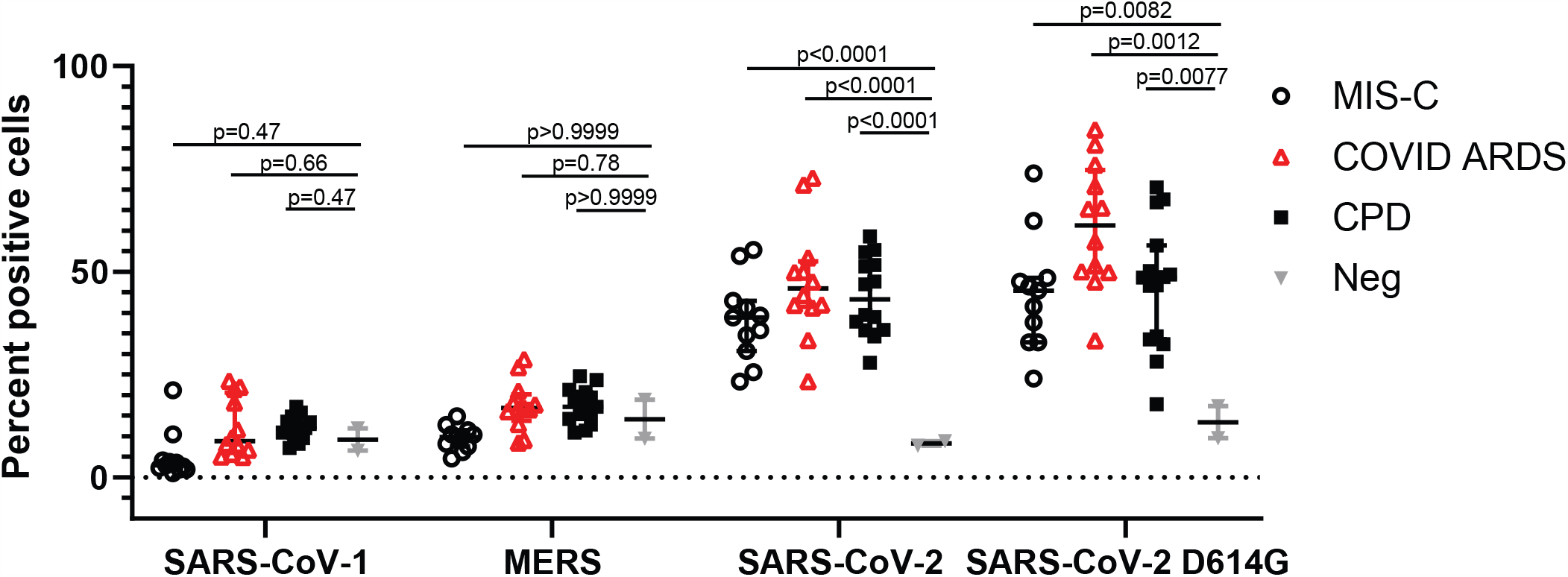
Patient anti-S IgG binds SARS-CoV-2 S protein but not SARS-CoV-1 and MERS S protein. HEK-293T cells were transfected with S protein and a common variant from the indicated coronaviruses. The transfected cells were then incubated with human plasma from the indicated study groups and bound human IgG was detected using fluorescently tagged protein G (see methods). Shown are the percentage of the S protein transfected cells that are positive for bound human IgG in each patient group: MIS-C, white circles, n=15; COVID ARDS, red triangles, n=14; CPD, black squares, n=19; and control plasma from pre-pandemic donors (grey triangles; Neg, n=8).

**Extended data figure 2.**
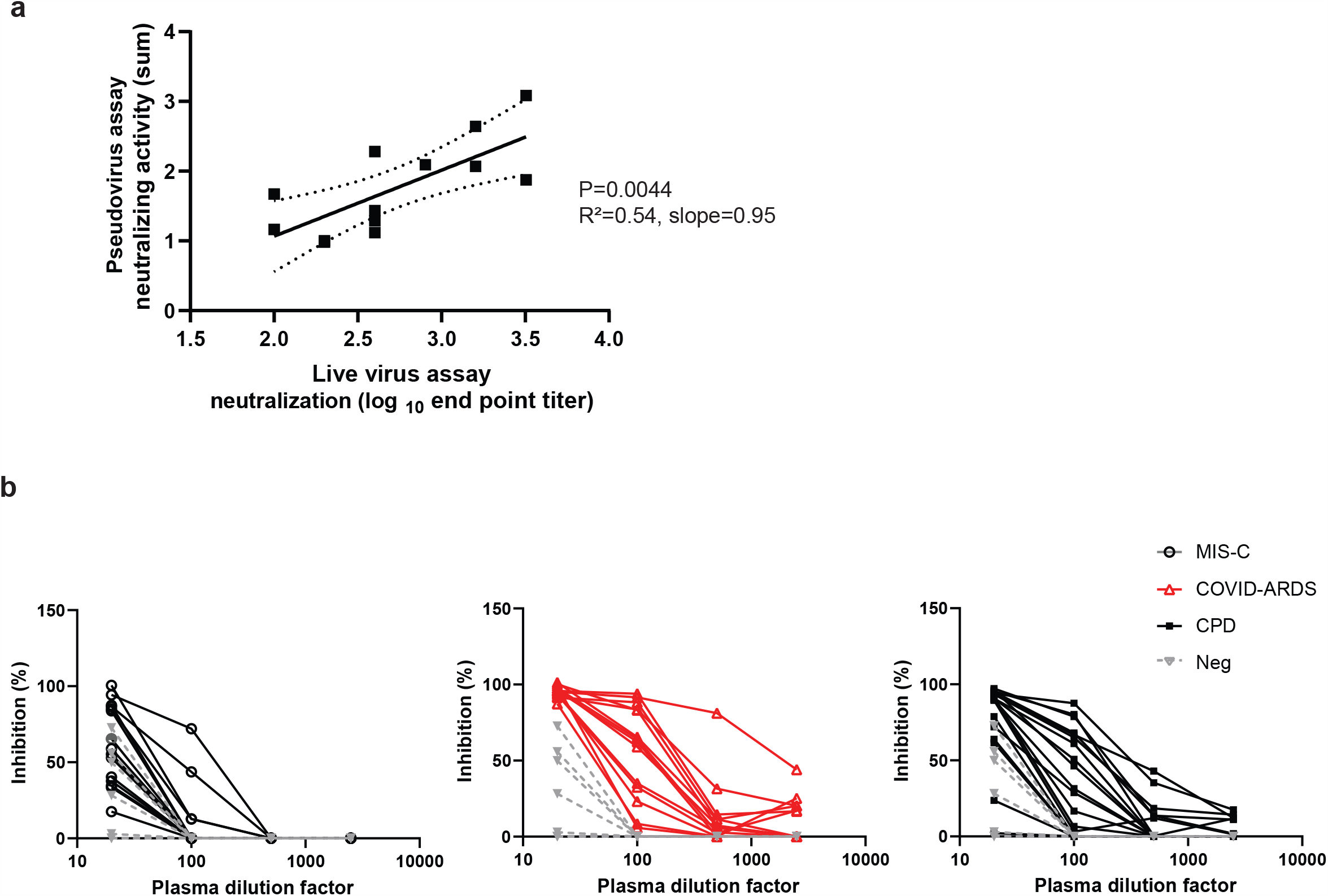
a, Plasma neutralizing activity in the pseudovirus assay was correlated with the end point titers in a live virus microneutralization assay based on inhibition of cytopathic effect (see methods). b, Using the results of the pseudovirus neutralization assay, the percent inhibition of S protein mediated multicycle replication was calculated for each patient plasma sample across all dilutions (see methods). Shown (b) are the percent inhibition values plotted against the plasma dilution factors in patients with MIS-C (white circles, n=15); patients with COVID ARDS (red triangles, n=14); convalescent plasma donors (CPD, black squares, n=19), and control plasma from pre-pandemic donors (grey triangles; Neg, n=8).

**Figure S1.**
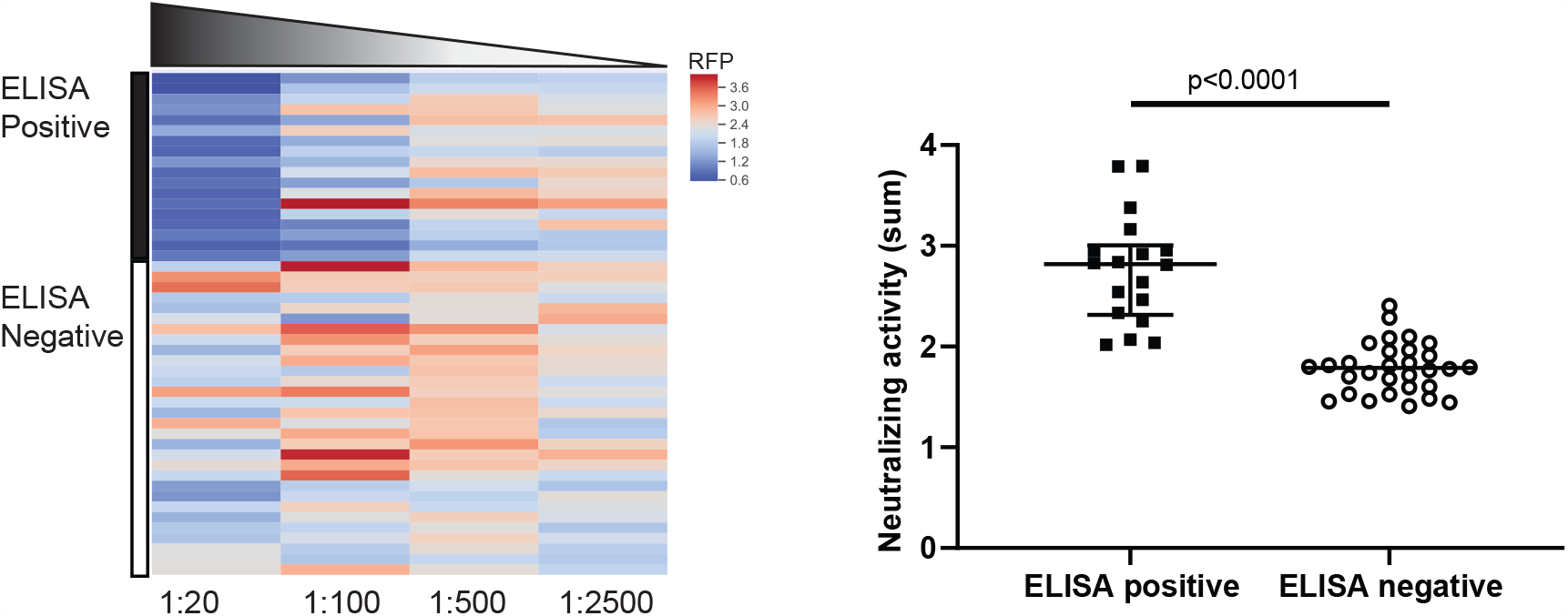
SARS-CoV-2 neutralizing activity in sera previousy tested for anti-SARS-CoV-2 S protein antibodies by ELISA. The pseudovirus-based assay was used to assess neutralizing activity against SARS-CoV-2 in sera that tested positive and negative for anti-S protein antibodies by ELISA. For each sample, neutralization activity is calculated based on inhibition of the pseudoviurs-driven RFP signal (see Figure 2 and methods). Shown is a heat map (left) of the normalized RFP signal across plasma titrations from de-identified patient samples that were sent to the clinical lab for ELISA-based SARS-CoV-2 antibody testing and were found to be positive for anti-S IgG by ELISA (black bar, n=18); and negative for anti-S IgG (white bar, n=30). The sum of the reciprocal values at the dilutions are shown (right) compiled from each of the groups. ±IQR. P value as calculated by Student’s t-test.

**Table S1:**
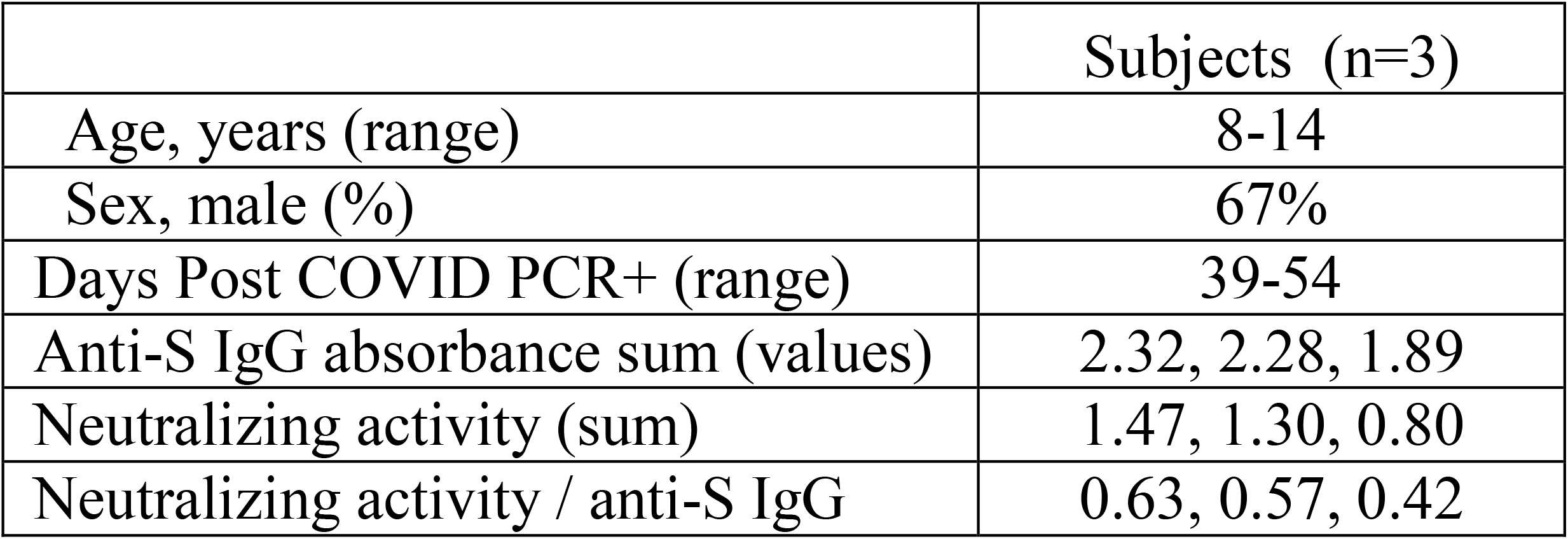
Anti-S antibodies in pediatric COVID patients without MIS-C

|  | Subjects (n=3) |
| --- | --- |
| Age, years (range) | 8-14 |
| Sex, male (%) | 67% |
| Days Post COVID PCR+ (range) | 39-54 |
| Anti-S IgG absorbance sum (values) | 2.32, 2.28, 1.89 |
| Neutralizing activity (sum) | 1.47, 1.30, 0.80 |
| Neutralizing activity / anti-S IgG | 0.63, 0.57, 0.42 |

